# Is Australia ready for the rollout of amyloid-targeting therapies for Alzheimer’s disease? Results from a national survey characterising current infrastructure capability, workforce and training needs of memory and cognition clinics

**DOI:** 10.1101/2024.07.05.24309974

**Authors:** Johannes C. Michaelian, Christopher C. Rowe, Susan E. Kurrle, Constance Dimity Pond, Michael Woodward, Sharon L. Naismith

## Abstract

**Background:** New amyloid-targeting monoclonal antibody (mAb) therapies for Alzheimer’s disease (AD) are currently under review by the Therapeutic Goods Administration for use in Australia.

**Aims:** To determine the infrastructure, workforce and training needs of Australian memory and cognition clinics to characterise health system preparedness for amyloid-targeting mAb therapies for AD.

**Methods:** A national, cross-sectional online survey of medical specialists was conducted.

**Results:** Thirty medical specialists (Geriatricians, n=23; Psychiatrists, n=4; Neurologists, n=3) from 30 different clinics participated (public, 76.7%; private, 23.3%), including metropolitan (73.3%), regional (20.0%) and rural (6.7%) areas. On average, clinics reported assessing 5.4 (SD=3.2) new patients per week, of which 2.4 (range: 0-5) were considered to have Mild Cognitive Impairment (MCI). Only 40% of clinics use biomarkers to assess whether patients with MCI have AD, and 55% have intravenous infusion capability. While the majority of clinicians were confident in their knowledge of mAbs, only 33% felt confident in using these. Identified impediments to clinical implementation included a) lack of real-world experience; b) lack of current Models of Care and appropriate use guidelines; c) current clinic set-up; and d) information about safety.

**Conclusions:** Australia’s health system preparedness for amyloid-targeting mAb therapies will require further investment in infrastructure, equity of access, clinician training and support. Long wait-times already impact access to clinics, and with the forecast rise in MCI and dementia cases, services will need to be expanded; while appropriate models of care and clear and efficient inter-sector health pathways will be needed to prepare for the use of mAbs.

## INTRODUCTION

Until recently, approved therapies for Alzheimer’s disease (AD) have been limited to symptomatic treatments that do not alter the underlying mechanisms of disease^1^. However, a new era of amyloid-targeting therapies for mild cognitive impairment (MCI) due to AD and early AD is imminent, spurred by breakthrough developments in the use of monoclonal antibodies (mAbs)^2^. These therapies stimulate the body’s own immune system to remove neurotoxic forms of amyloid-beta (Aβ). In the past two years, clinical trials of two agents, lecanemab and donanemab, have shown that these drugs slow cognitive decline over 18-months, with concomitant benefits on disease biomarkers and quality of life^3, 4^.

In Australia, these agents are currently before the Therapeutic Goods Administration (TGA) for regulatory approval. While lecanemab has gained approval in the US, Japan and China, such that the international community is already underway in their preparedness and clinical implementation^5–7^, there is a need to consider Australia’s health system readiness for such therapies. Notably, several investigations will be required to confirm a prospective patient’s eligibility, in addition to regular infusions and the close safety monitoring of the patient for amyloid-related imaging abnormalities (ARIA).

In this study, we therefore aimed to conduct a national survey of medical specialists involved in the assessment of dementia and cognitive decline in order to determine:

1. The infrastructure capability and additional needs to support amyloid-targeting mAb therapies for AD;
2. The clinical workforce capability needed to deliver an amyloid-targeting mAb therapy; and
3. Training needs/knowledge including skills gaps to adequately deliver treatment of an amyloid-targeting mAb therapy.

## MATERIALS AND METHODS

### Sample and Setting

The sampling frame for the survey included geriatricians, neurologists, or old age psychiatrists employed in a specialist assessment clinic for dementia and cognitive decline (i.e., the clinic did not have to identify as a Memory Clinic, Memory and Cognition Clinic or CDAMS). Several recruitment strategies were employed. First, a single email invitation to participate in the survey was sent to the official contacts of clinics listed on the Australian Dementia Network (ADNeT) *Memory Clinic or Cognition Decline Assessment Service Online Finder Tool*^8^. Three reminder emails were sent. In addition, the survey was advertised via social media, such as the ADNeT LinkedIn account, and within professional networks and organisations (i.e., Australian & New Zealand Society for Geriatric Medicine). The survey was open from the 7^th^ of September 2023 until the 7^th^ of February 2024.

### Survey and Procedure

The survey was delivered in Qualtrics^9^. Participants were given the option to complete the survey online or via phone-call with the study coordinator. The conduct of this study was approved by the University of Sydney’s Human Research Ethics Committee (Approval Number: 2023/480). All potential respondents provided informed consent, electronically or verbally via phone-call with the study coordinator. All study methods were conducted in compliance with the Helsinki Declaration.

The survey comprised 5 sections: 1) Current clinical landscape of diagnosis; 2) Current treatment journey/workforce; 3) Treatment; 4) Logistics/capacity; and 5) Knowledge. A mix of structured (e.g., “agree/disagree”, “yes/no”) including multiple-choice “checkbox” or single-choice “radio buttons” answer options were employed, in addition to unstructured (e.g., free-text box) answer options.

### Data analysis

All survey responses were recorded and initially saved in Qualtrics^9^. Statistical analysis was conducted in SPSS Version 27 (SPSS Inc., IBM Corp. in Armonk, NY, USA). Descriptive analyses (i.e., frequencies and percentages) were performed for outcome measures. Missing or ‘unable to comment’ responses were recorded, and the total number of ‘valid responses’ became the denominator for that item.

## RESULTS

### Respondents

As shown in Table 1, responses were received from 30 medical specialists (Geriatricians, n = 23; Psychiatrists, n = 4; Neurologists, n = 3) across 30 different clinics including 23 publicly funded clinics (76.7%) and 7 private clinics (23.3%), of which 2 were clinics embedded within universities. The majority were in metropolitan areas (n = 22, 73.3%) followed by regional (n = 6, 20.0%) and rural (n = 2, 6.7%) areas. While there was representation from all Australian states/territories except for the Australian Capital Territory (ACT), New South Wales (n = 10, 33%) and Victoria (n = 10, 33%) accounted for two-thirds of the respondents in proportion to population and workforce (Figure 1). Approximately two-thirds of clinics (n = 19, 63.3%) offered telehealth and 16.7% (n = 5) offered a roving/mobile service.

**Table 1:**
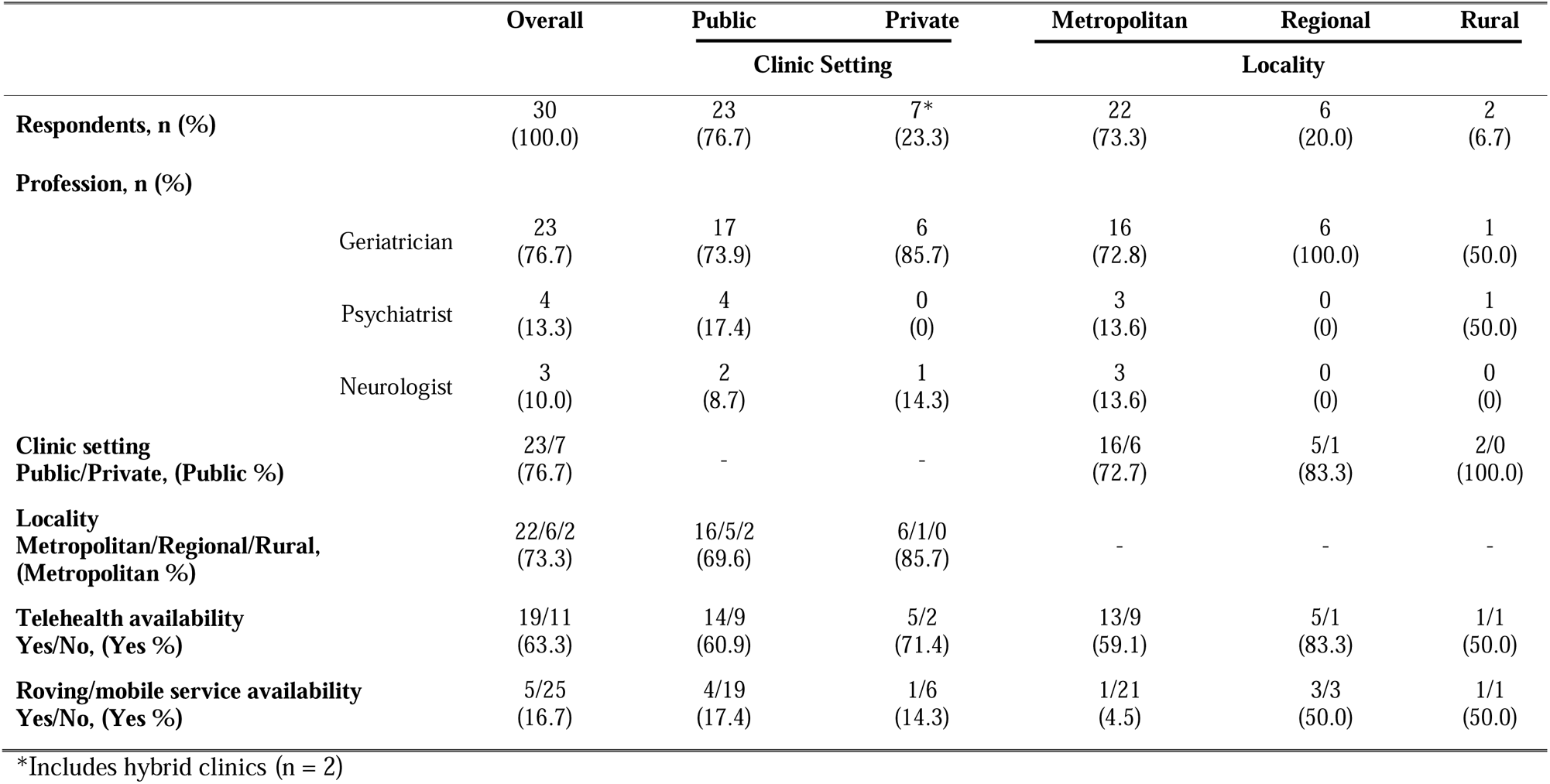
Characteristics of survey respondents.

**Figure 1:**
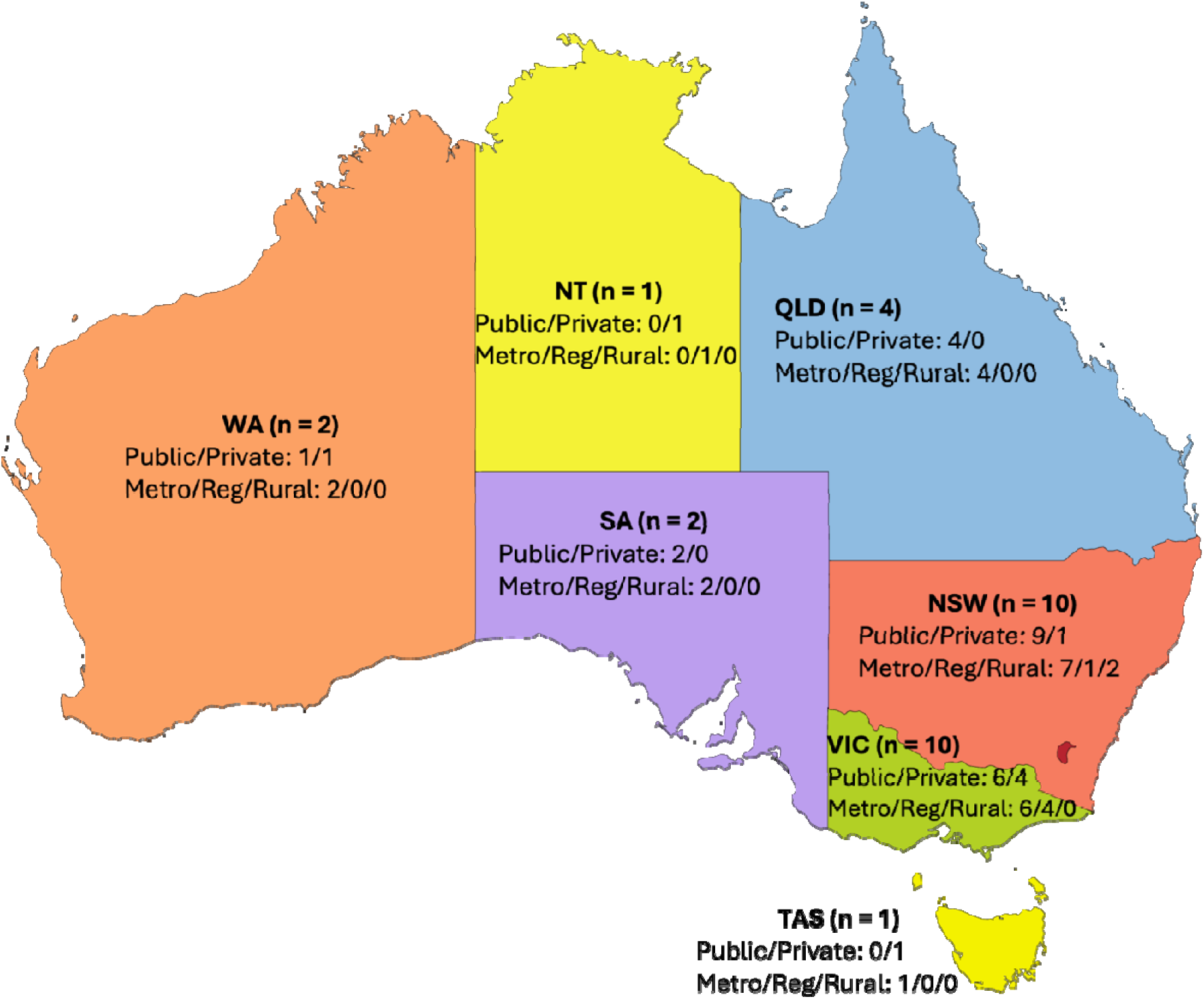
Survey representation across Australian states and territories for public and private clinics located in metropolitan, regional and rural regions. NSW = New South Wales, NT = Northern Territory, QLD = Queensland, SA = South Australia, TAS = Tasmania; VIC = Victoria, WA = Western Australia

### Current clinical landscape of diagnosis

#### Referral pathways and number of patients assessed

The largest proportion of patient referrals were from a General Practitioner (GP) (78.3%), followed by a medical specialist (15.9%). Less than 10% of clinics received referrals from Allied Health professionals, Aged Care Assessment Teams (ACAT) and self-referrals. On average, clinics reported seeing 5.4 (SD = 3.2, range = 1 to 15) new patients per week, as well as 9.4 follow-up patients per week (SD = 6.7). Approximately 60% and 31% of patients travelled between 5-20 kilometres or 20-40 kilometres for their assessment, respectively.

#### Clinical assessment of Mild Cognitive Impairment (MCI) and MCI due to Alzheimer’s disease

Almost all clinics (n = 29, 96.7%) assessed people for having MCI, and estimated an average of 2.4 patients would meet MCI clinical criteria (range: 0-5). However, only 40% (n = 12) of clinics reported using imaging *biomarkers* (e.g., FDG-PET, Aβ- and/or tau-PET) in their diagnostic work-up. A minority (n = 8, 26.7%) reported that if needed, they could refer patients for confirmatory Aβ- and/or tau-PET imaging, but only one clinic performed this routinely (>70% patients). Indeed, 60% (n = 18) of clinics reported that cerebrospinal fluid (CSF) sampling to determine a patient’s Aβ status was never / not available for testing, and a further 30% (n = 9) would rarely refer their patients for testing. Similarly, more than 60% (n = 19) of clinics reported that ApoE genotyping was never / not available for testing.

When clinicians were asked to describe current challenges and barriers towards the implementation of an early diagnosis of MCI due to AD, as illustrated in Figure 2, four main themes emerged. These were “access and availability” constraints as well as the “out of pocket expense to patients” likely to be incurred for confirmatory biomarker investigations (i.e., Aβ- and/or tau-PET imaging). Clinicians also reported that were a referral to be made for imaging biomarker investigation, the “current wait-times” inhibited receiving results to assist in the diagnostic work-up. Moreover, clinicians described that their patients were not presenting “early in the trajectory of their illness” (i.e., in later dementia stages).

**Figure 2:**
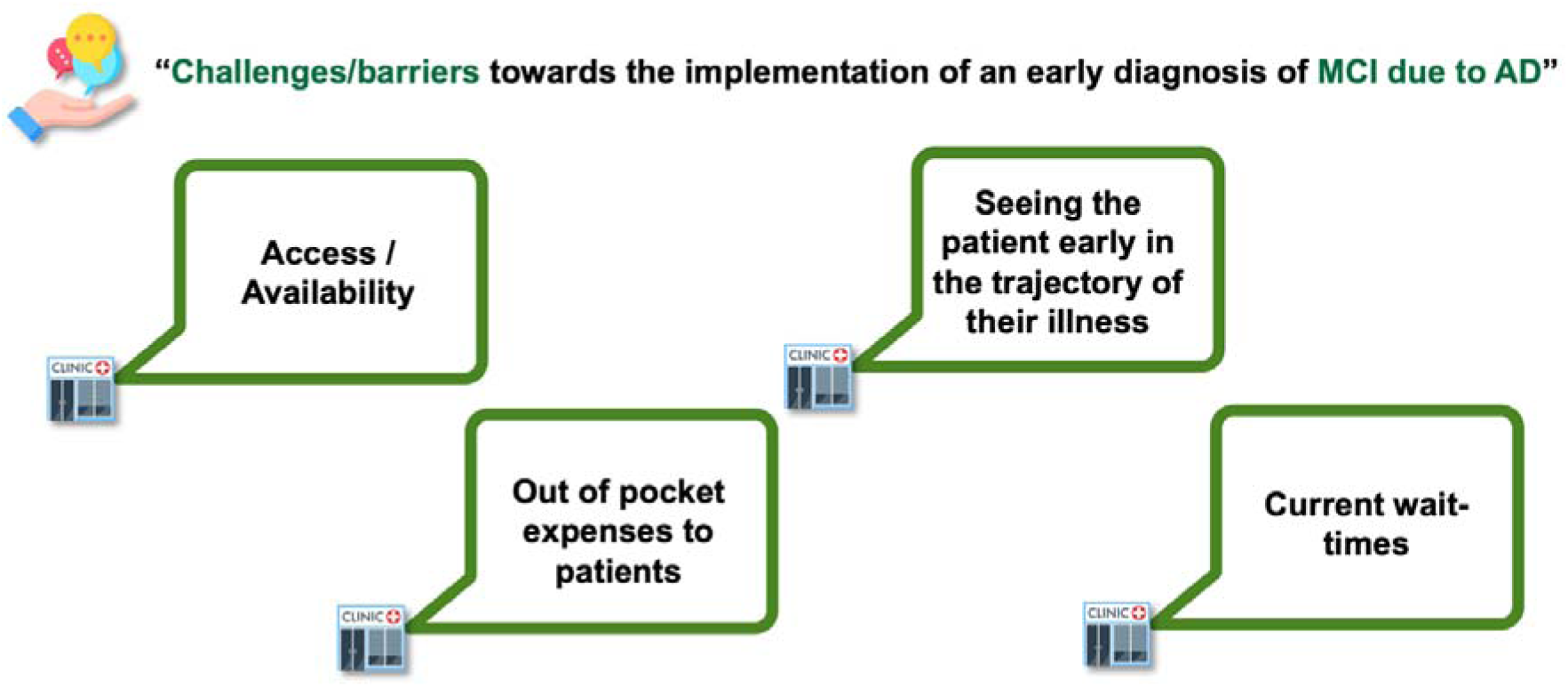
Challenges/barriers most commonly inhibiting the implementation of an early diagnosis of mild cognitive impairment (MCI) due to Alzheimer’s disease (AD) clinically

#### Magnetic Resonance and Positron Emission Tomography imaging investigations

The majority of clinicians (n = 26, 86.7%) indicated that patients with MCI or clinical AD would be referred for a brain MRI scan if appropriate, and on average estimated that 59.0% (SD = 33.8) of new patients would be referred for an MRI, a result which remained consistent between public (n = 19, 58.3% ± 34.4% [mean ± SD]) and private clinics (n = 7, 60.9% ± 34.5% [mean ± SD]). Only a minority of clinicians (n = 8) refer those with MCI or suspected AD dementia for confirmatory Aβ and/or tau PET imaging. Consequently, approximately a quarter of new patients seen within a clinic are referred for a brain PET scan (n = 7, 26.6% ± 29.1), including referral for FDG-PET. Table 2 reports the wait-times (in days) for brain MRI and PET scans. Almost two-thirds (n = 19, 63.3%) of clinics estimated that their service had on-site capacity to conduct brain MRI scans for diagnosis and monitoring, and the majority reported that it was likely (n = 16, 61.5%) to extremely likely (n = 7, 26.9%) that the same scanner could be used for multiple scans (i.e., for ARIA safety monitoring).

**Table 2:**
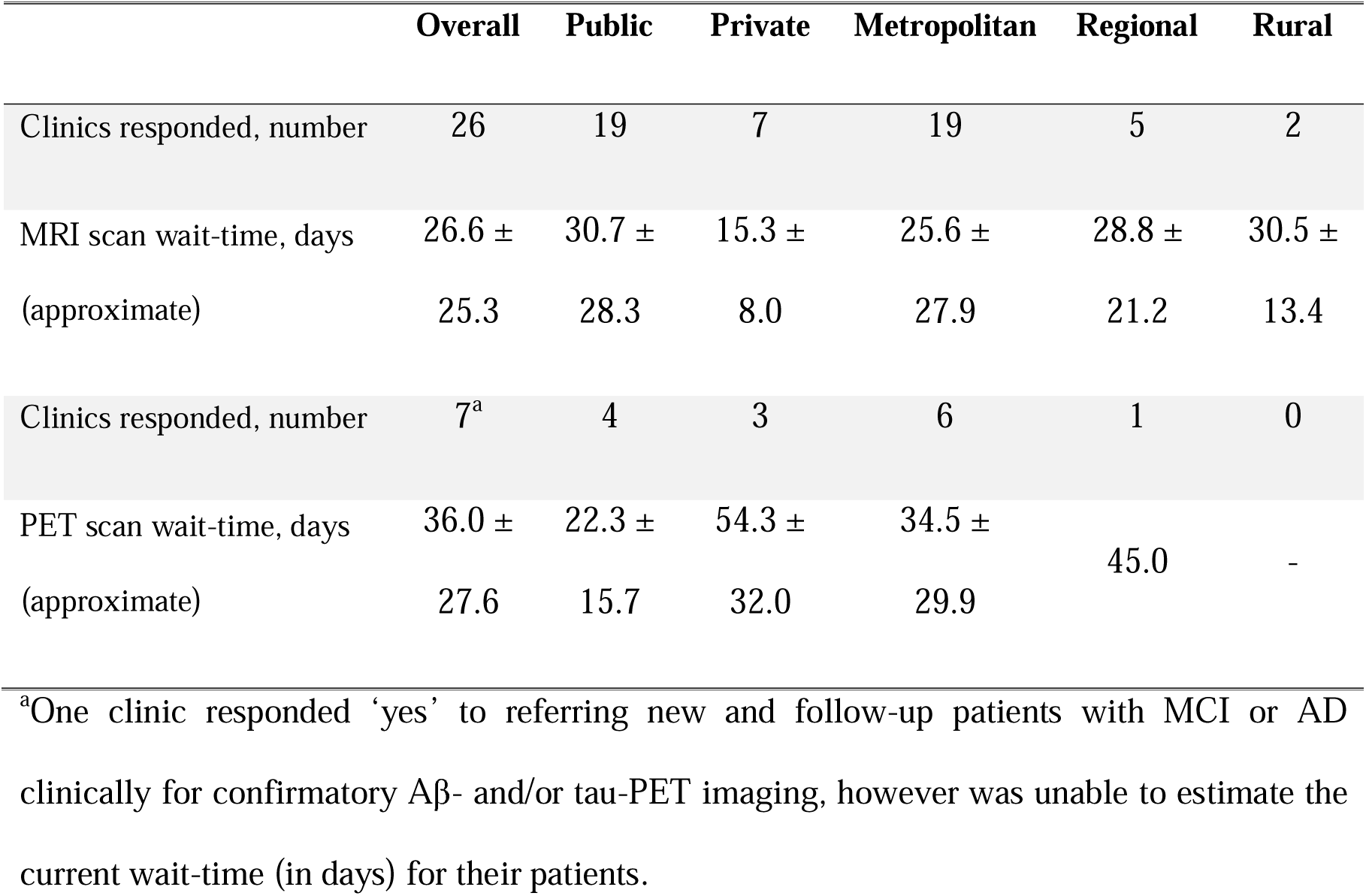
Reported approximate wait-times for a brain MRI or PET scan across clinics surveyed.

### Current treatment journey/workforce

The majority of clinics (n = 21, 70%) employed multi-disciplinary teams for diagnosis and 66.7% (n = 20) provided some form of post-diagnostic support for MCI and AD, largely comprising one feedback session (n = 14, 70%), referral to Dementia Australia (n = 17, 85%) and dementia support websites (n = 16, 85%). For patients with MCI due to AD, 66.7% (n = 20) of clinicians did not prescribe pharmacological treatments, 33.3% (n = 10) were prescribing an off-label treatment, of which acetyl-cholinesterase inhibitors were the most common (n = 9). Only 36.7% (n = 11) of clinicians currently provide training to local GPs in the area of MCI and/or dementia assessment and management.

### Treatment

When clinicians were asked to indicate appropriate targets for treatment for AD, the majority indicated that Aβ (n = 25, 83.3%) and addressing vascular risk factors (n = 26, 86.7%) should be targeted, while only half (n = 15) reported that phosphorylated-tau (i.e., neurofibrillary tangles) should be a treatment target. In addition, management of other / modifiable risk factors were identified as key targets (n = 25, 83.3%), with exercise/physical inactivity, inflammation and sleep commonly mentioned. Regarding the impact of treatments, as illustrated in Figure 3, all clinicians reported that it was very important (n = 14, 48.3%) to important (n = 13, 44.8%) that any treatment for AD has a statistically significant impact on activities of daily living (ADL). Moreover, as to be expected for any treatment, real-world evidence of clinical efficacy was considered to be very important (n = 23, 76.7%) alongside real-world evidence of safety (n = 26, 86.7%). Greater than 96% of clinicians also considered cognitive abilities, functional ability and dependence, behavioural and neuropsychiatric symptoms as well as patient quality of life to be important markers to gauge the efficacy of any new treatment for AD.

**Figure 3:**
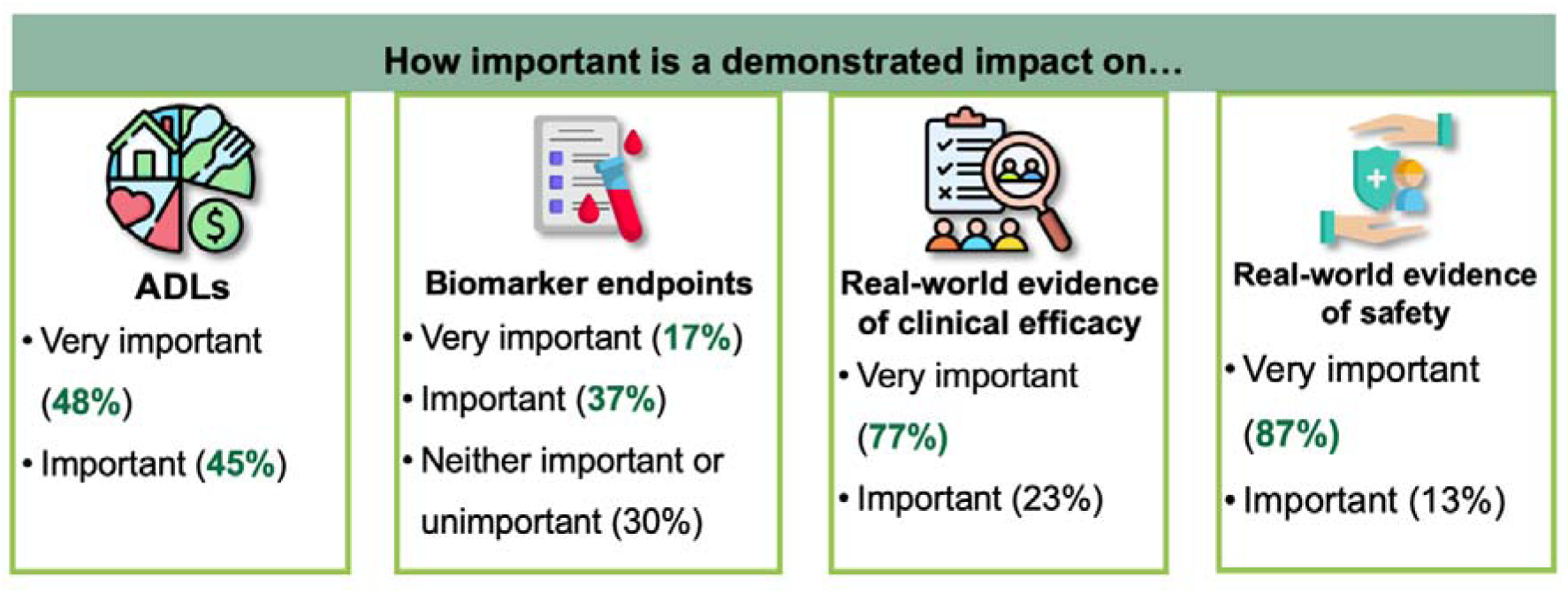
Importance of demonstrated impact for any new treatment for Alzheimer’s disease

As summarised in Figure 4, clinicians reported that for any treatment for AD a clinically meaningful 6-12 month outcome would either ‘improve’ or ‘maintain’ activities of daily living (ADLs), independence and cognitive abilities as well as ‘slow down’ or ‘stabilise’ cognition, function or AD progression.

**Figure 4:**
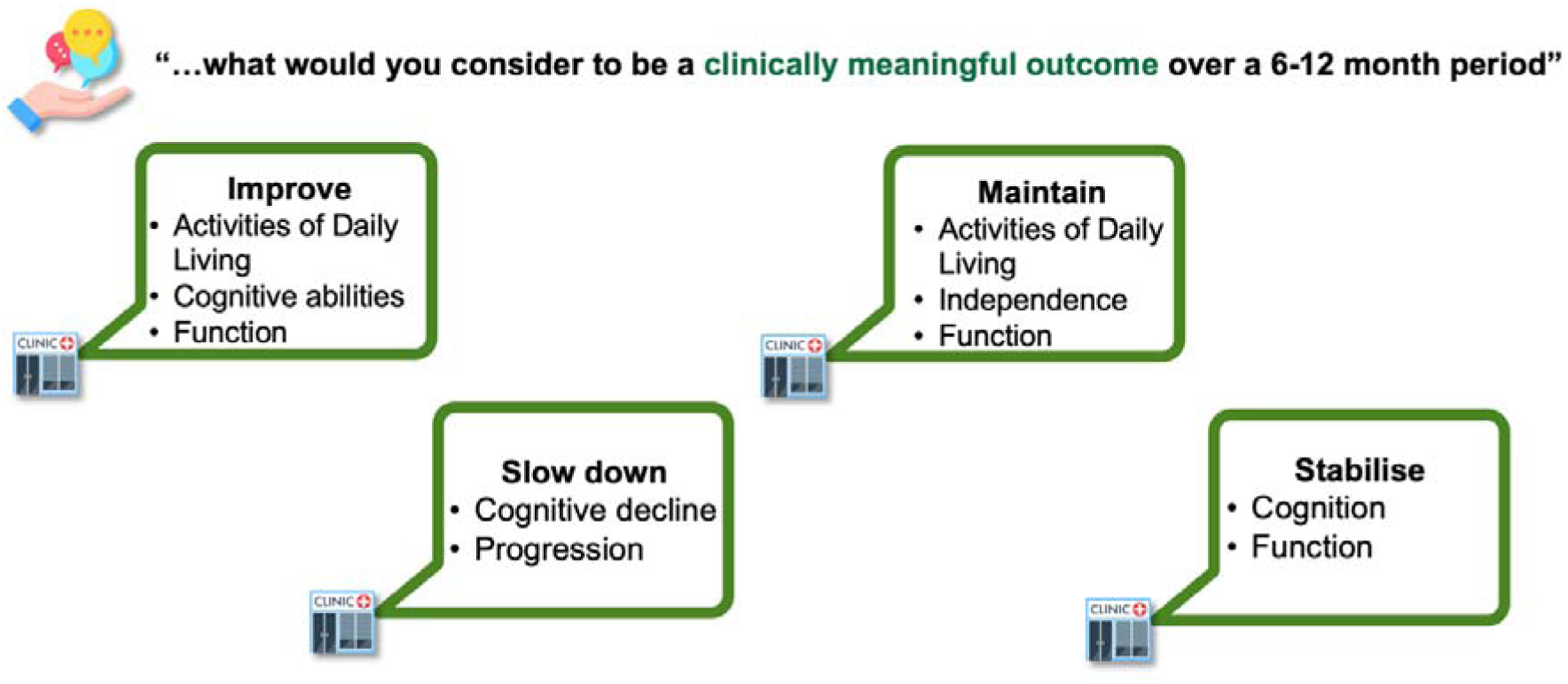
Clinically meaningful outcomes considered by to be important over a 6-12 month period for any new treatment for Alzheimer’s disease

### Logistics/capacity

Logistically, two-thirds of clinics (n = 20, 66.7%) reported that a drug-specific refrigerator (2-8 °C) was available for storage, and of these clinics demonstrating current capacity, over sixty percent (n = 11, 61.1%) could accommodate increased demand for an amyloid-targeting mAb therapy requiring cold storage. More than half of responding clinics (n = 16, 55.2%) reported that their service did not currently have the required resources to facilitate intravenous (IV) administration on-site using an infusion chair for mAb therapies requiring regular administration (i.e., fortnightly), but of this number close to half (n = 7, 43.8%) would utilise a home infusion service to facilitate IV administration.

More than half of clinics (n = 17, 56.7%) reported current on-site nursing and administrative support for follow-up/coordination of patient appointments and infusions. In terms of clinician confidence in prescribing and administering an amyloid-targeting mAb therapy requiring IV administration, only 33% of clinicians were ‘very confident’ (n = 6) or ‘confident’ (n = 4) in IV administration, with the latter needing refresher training and support. A further 13.3% (n = 4) were ‘somewhat confident’ but noted they would need training, while 53.3% endorsed items indicating they were ‘not at all confident’ and were *not* interested (n = 7) or required support if they were interested (n = 9). For the latter, clinician support would include an appropriate nursing workforce (n = 9, 100%), training (n = 9, 100%) and infrastructure (n = 8, 100%). Clinicians also overwhelmingly reported that they (over their patient’s GP) would prefer to review their patient receiving a mAb therapy, on a monthly (n = 10, 43.5%) to quarterly (n = 11, 47.8%) basis. Lastly, when clinicians were asked to share any comments about IV administration, comments centred around staffing needs, location, resources/infrastructure needs as well as capacity (Figure 5).

**Figure 5:**
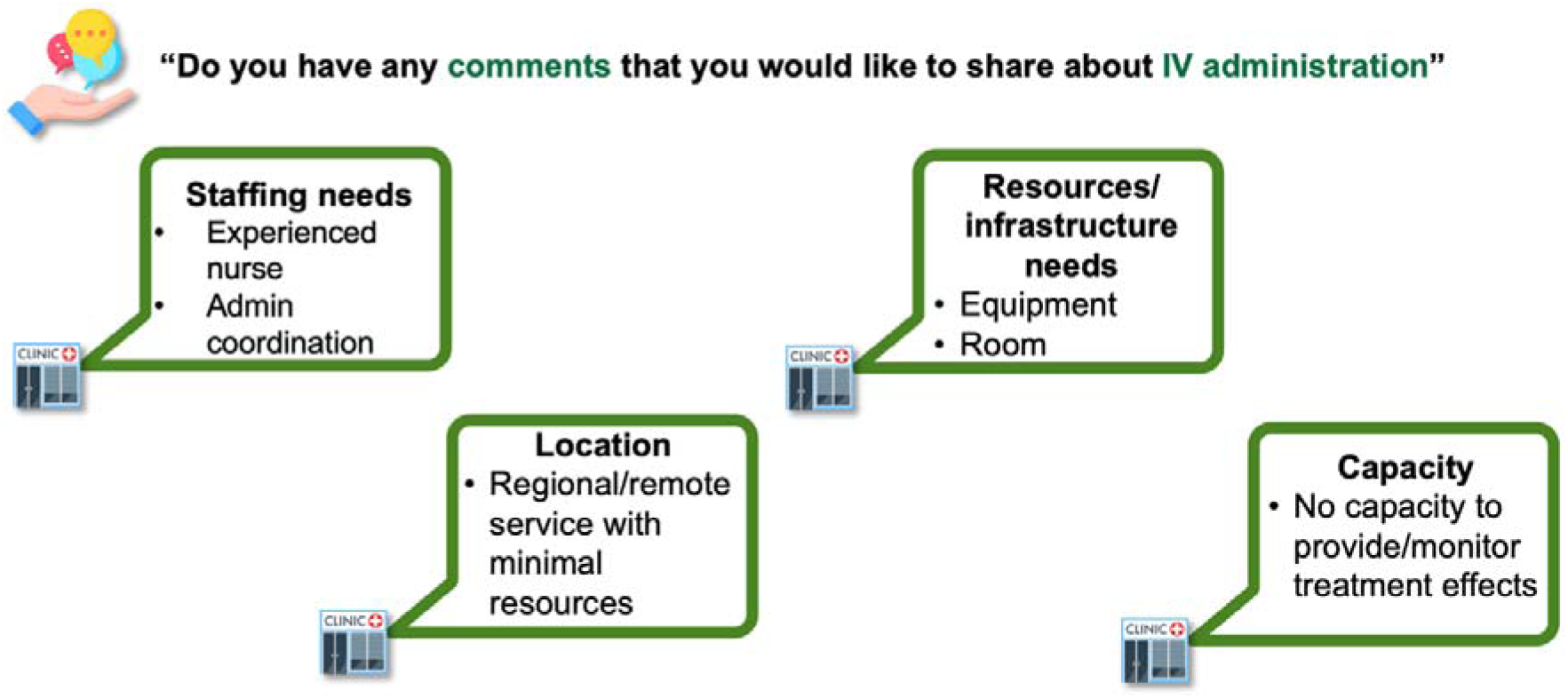
Overall comments received by clinicians around the IV administration of an amyloid-targeting mAb therapy within their clinic/service.

### Knowledge

Despite the majority of clinicians reporting that they have not been involved in a recent clinical trial of an amyloid-targeting therapy (n = 21, 72.4%), the majority (80%) reported *confidence* around their knowledge on the mechanisms of action of a mAb against Aβ: ‘somewhat’ (n = 12, 40%), ‘quite’ (n = 9, 30%) and ‘extremely’ (n = 3, 10%) confident. Only three clinicians reported being ‘not at all confident’ (10%). Indeed, the majority of clinicians (80%) also reported *confidence* around their knowledge on the clinical trial outcomes of an amyloid-targeting therapy: ‘somewhat’ (n = 12, 40%), ‘quite’ (n = 7, 23.3%) and ‘extremely’ (n = 5, 16.7%) confident.

On the whole, clinicians reported a desire to receive information about patient support programs (n = 27, 93.1%), reimbursement/insurance coverage (n = 25, 86.2%) and around the detection/management of ARIA (n = 18, 62.1%) if/when these become available. In terms of receiving training around amyloid-targeting mAb therapies for AD, a clinician’s most preferred method was webinars/online demonstration (n = 16, 57.1%) followed by on-site hands-on demonstration (n = 9, 32.1%) and external seminar/workshop (n = 3, 10.7%).

Lastly, clinicians were asked to comment on what they perceived to be the greatest knowledge gaps towards the implementation of an amyloid-targeting therapy in their service. As illustrated in the Figure 6, responses covered several different themes, for example:

- Lack of real-world experience; suggesting that there is a need to understand how treatment of a patient living with AD can be translated from the clinical trial environment to a real-world clinical setting in Australia;
- Lack of current Models of Care, appropriate use criteria and protocols around infusion and therapy monitoring;
- Current clinic set-up; such that clinics are not set up to administer an amyloid-targeting therapy on-site with an infusion chair; and
- Information about safety; identifying and managing ARIA as well as overall safety monitoring.

**Figure 6:**
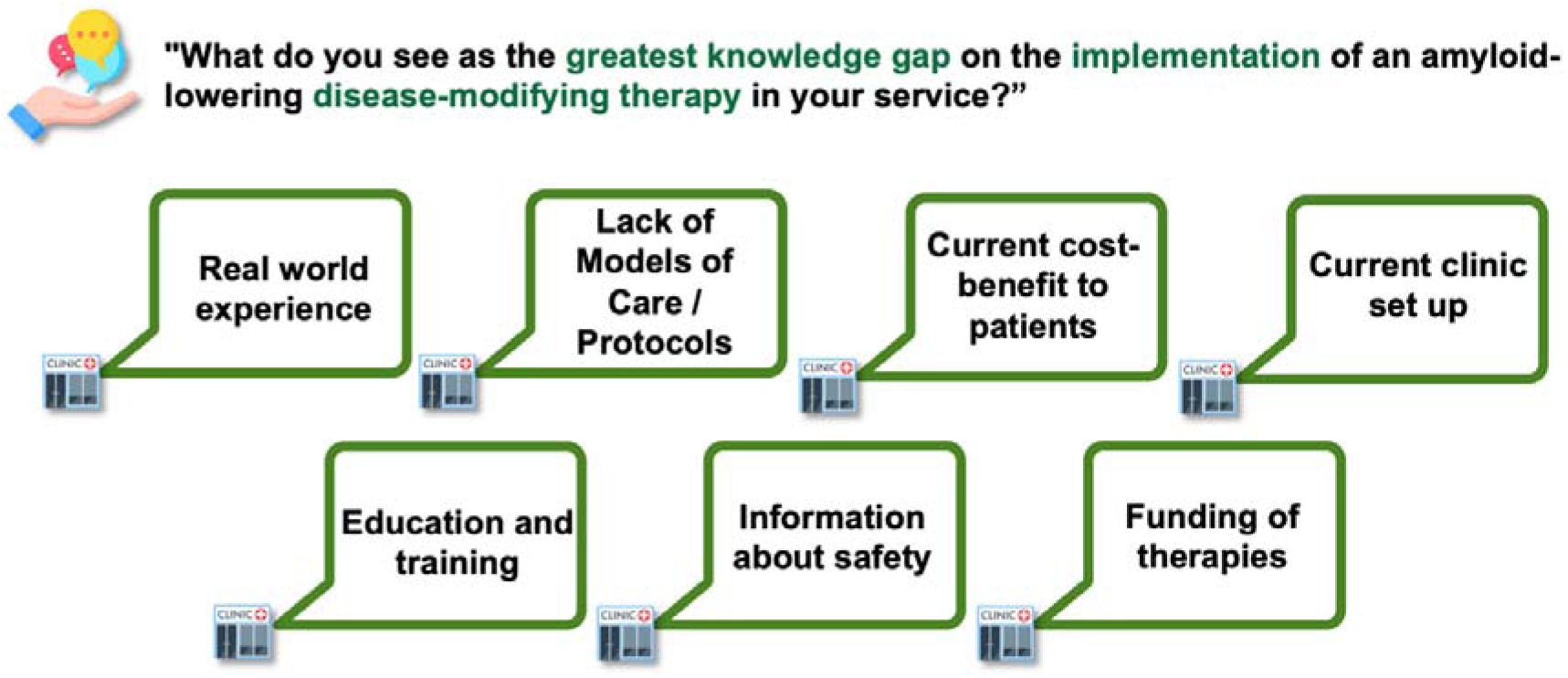
Overall themes to emerge when clinicians were asked to comment on the ‘greatest knowledge gaps’ surrounding the clinical implementation of an amyloid-targeting mAb therapy within their service

## DISCUSSION

This study overall shows that if amyloid-targeting mAb therapies were to become available in Australia, improvement of current infrastructure capability, workforce and training will be required. Findings affirm that while clinicians were confident in their knowledge of the mechanisms of action of amyloid-targeting mAbs, further data on real-world use as well as cognition and activities of daily living outcomes was anticipated. The survey also reveals that medical specialists perceive other / modifiable risk factors should be targets for treatment, including exercise/physical inactivity, inflammation and sleep. To support this, other forms of post-diagnostic support around cognitive interventions, carer support programs, and care navigators will be vital, within and between primary care^10^. Indeed, comprehensive management options will be especially important in light of international figures showing that only a small proportion (lecanemab, 8%; aducanumab, 5%) of patients presenting to a memory and cognition clinic would be eligible for an amyloid-targeting mAb therapy if the criteria used in clinical trials were applied to routine clinical practice^11^.

In terms of the health service preparedness, importantly, this study shows that Australian memory and cognition clinics are limited to assessing around 5.4 patients per week, of which approximately two cases may have MCI. This estimate is concordant with our prior national surveys^12, 13^, and with ADNeT Clinical Quality Registry data which shows that ∼32% of new memory and cognition clinic cases meet *clinical* criteria for MCI (i.e., not necessarily MCI due to AD)^14^. Of concern, however, most gold-standard memory and cognition clinics only operate 1-2 days per week, and may only be able to service around 5% of MCI/dementia cases, demonstrating substantial unmet need^15^. Indeed, recent estimates suggest that almost 12,000 patients per year may attend a public memory and cognition clinic^15^. Extrapolating from these figures, and assuming clinics had the relevant infrastructure, capability and workforce needs in place, approximately 1000 patients from the first year attending a public clinic could be eligible to receive lecanemab (i.e., 8% of 12,000 patients eligible)^11^ when running at current capacity, with prescriptions also likely dependent on Pharmaceutical Benefits Advisory Committee (PBAC) and Medical Services Advisory Committee (MSAC) approvals in addition to TGA approval. However, given there will be approximately 250,000 new MCI and dementia cases per year in those aged over 65 years^15^, up to 20,000 Australians could be suitable for lecanemab (8% of 250,000), noting that co-existing chronic diseases and neuroimaging findings are most likely to warrant a patient ineligible^11^. These figures, while only an approximation, suggest that public services could meet the demands for only 5% (i.e., 1000/20,000) of suitable cases. Thus, if lecanemab and donanemab were to be available and receive government subsidy in Australia, there would need to be substantial investment in expansion of public sector memory and cognition clinics, as well as key considerations and support of private sector models. Notably, there would also need to be increased access and funding for Aβ-PET imaging or lumbar puncture for CSF biomarker assays, infusion capabilities, nursing workforce and administration support, as well as frequent neuroradiological reviews. In the future, with the introduction of new blood (plasma) biomarkers for AD^16^, screening would become more effective and better integrated with primary care, and trials are underway to test the use of these plasma biomarkers in Australian memory and cognition clinics (e.g., ACTRN12622000515796) and work has begun to scope how these could best be utilised in primary care settings.

This study also highlighted that workforce training would be required. Specifically, clinicians would require support and training in the IV administrations of an mAb therapy as well as support for patients, and reimbursement/insurance coverage and training around ARIA detection/management and acute infusion reactions.

One of the major findings of the survey to emerge was the need for clear and comprehensive Models of Care and appropriate use guidelines. This is important as clinicians noted a lack of real-world experience in the use of an amyloid-targeting mAb therapy, particularly within context of the Australian health care system. Such guidelines will be highly important in understanding the frequency of MRI monitoring to detect ARIA, especially as ARIA cases with cerebral oedema (ARIA-E) and microhemorrhage/hemosiderosis (ARIA-H) are common for both donanemab (ARIA-E 24.0%; ARIA-H 31.4%) and lecanemab (ARIA-E 12.6%; ARIA-H 17.3%)^3, 4^. Given that use of the same MRI scanner across multiple timepoints is recommended to attenuate artifacts either due to different scanning protocols and/or magnet strength^17^, it was a positive finding that the vast majority of clinics indicated that it was likely to extremely likely that the same MRI scanner could be used across multiple scans. Nevertheless, for GPs and emergency specialists who may be involved in the routine or inadvertent management and care of a patient receiving an amyloid-targeting mAb therapy, further education and training is needed for them to discern between therapy-related ARIA and ischaemic stroke. Monitoring of real-world outcomes would also be important and may occur alongside the Australian Dementia Network Clinical Quality Registry for participating clinicians^14^. Given that two-thirds of medical specialists surveyed are not currently providing training to GPs in the area of MCI and/or dementia, there will also be a need to provide critical education and support to GPs and practice nurses who will be fielding enquiries, seeking specialist input and advice as well as managing potential questions and side-effects that may arise during the course of treatment.

### Limitations

The present study has limitations. While there was broad national representation (except for the ACT), not all memory and cognition clinics previously identified by ADNeT participated in the survey. The nominated medical specialist completing the survey may have also interpreted and responded to questions specific to their clinical knowledge and practice, which may not be representative of other clinicians within that practice.

## Conclusion

Overall, the successful clinical implementation of amyloid-targeting mAb therapies for AD will rely on expanding memory and cognition clinic capacity, improving infrastructure capability, and addressing workforce and training needs. There will also be a need to implement appropriate use guidelines and support primary care in training and support; in addition to models of care and health pathways, particularly for those patients that are not eligible - which is estimated to be around 92% of MCI and early AD patients. It is expected that the real-world rollout of such therapies, may occur slowly, within key metropolitan centres, due to the infrastructure and workforce limitations in regional and rural areas. However eventually, with service expansion and developments in screening (e.g., plasma biomarkers for AD), and optimised methods of drug delivery (e.g., subcutaneous administration), equitable access for all Australians seeking treatment would be expected.

## Data Availability

The data for this study will not be shared, as we do not have permission from the participants or ethics approval to do so.

## Acknowledgements

We are grateful to all Australian medical specialists who participated in our survey. Additionally, we would like to thank the Australia and New Zealand Society for Geriatric Medicine (ANZSGM) for distributing our survey to their members. We also want to thank all members of the Australian Dementia Network (ADNeT) for their support and ongoing collaboration. This study was supported by an investigator-initiated research grant from Eisai Australia and we are grateful for their support.

